# Genomic investigation reveals long-term local and regional circulation of dengue virus associated with recent outbreaks in Kenya

**DOI:** 10.1101/2025.06.05.25329058

**Authors:** Solomon K. Langat, Sheila Kageha, Victor Jeza, Genay Pilarowski, Albert Nyunja, Paul E. Oluniyi, Juliana Gil, Nelly Ogada, Lewis Gande, Jane Thiiru, Seth Okeyo, Pascah Bulia, Jael Amugongo, Victor Ofula, Francis Mutuku, Cristina M. Tato, Samoel Khamadi, Joel Lutomiah, Samson Limbaso

**Affiliations:** Centre for Virus Research, KEMRI, Nairobi, Kenya; Technical University of Mombasa, Mombasa, Kenya; Chan Zuckerberg Biohub, San Francisco, USA

**Author notes:** These authors contributed equally to this work (Solomon K. Langat, Sheila Kageha).

## Abstract

Dengue is an arbovirus disease caused by dengue virus. The virus has recently caused a global surge in cases, with Africa reporting over 250 million and 900 deaths since 2023. This study was conducted during the 2023 dengue outbreak in Mombasa, with the aim of understanding outbreak cases in the context of earlier cases and recent global epidemics. During the 2023 outbreak, 56 of 406 samples tested positive for DENV: including DENV-1, DENV-2, and DENV-3. The study generated 38 whole genome sequences, 24 from 2023 and 14 from cases identified between 2020 and 2025. Phylogenetic analysis showed DENV-2 exhibited patterns suggesting long-term local circulation, while DENV-1 and DENV-3 were more widespread regionally, and are linked to recent outbreaks in Eastern and Western Africa. These findings highlight the need for integrated strategies to curb dengue in Kenya, such as measures to disrupt local transmission as well as importation of new strains.

## Background

Dengue is an arbovirus disease caused by either one of the four serotypes of dengue virus; dengue virus 1-4 [1, 2]. Globally, dengue cases have increased substantially over the recent past, with more than 13 million cases and 8700 deaths reported in 2024 [3]. This was a marked increase from 2023, which saw a then-record 4.6 million cases being reported globally [4]. Africa has also documented an unprecedented increase in dengue cases since 2023. According to the World Health Organization (WHO), several African countries reported over a quarter million cumulative cases of dengue between 2023 and 2024 and approximately 900 deaths [5]. This multi-country outbreak of dengue affected 13 countries in West Africa and two countries in Eastern Africa including Ethiopia and Kenya [5]. However, the lack of sustained active surveillance of this disease in several African countries, especially those that border Ethiopia and Kenya could suggest an underestimation of the burden of dengue.

In Kenya, the first dengue associated outbreak was documented in the coastal town of Malindi in 1982 [17]. This was followed by a long period of relative quiescence. However, in the period between 2010 and 2014, there was a re-emergence of dengue in the country with multiple cases being reported in several counties [6, 7]. Following this outbreak, there have been almost yearly reports of dengue outbreaks, even though each of the subsequent outbreaks was mostly linked to one serotype [8, 9, 10]. Genomic analysis of viruses associated with the dengue outbreak reported in 2010 and subsequent years have found evidence that the virus was possibly introduced into Kenya from India in South Asia prior to 2010 [16]. The outbreaks between 2010 and 2014 were caused by three serotypes DENV 1-3, and it occurred in both the coastal region and the north eastern region of Kenya. However, subsequent outbreaks in 2017 and 2019 were associated mostly with dengue 2 and dengue 3 respectively [8, 9]. There was limited surveillance between 2020 and 2022, possibly due to the global SARS-CoV-2 pandemic where most of the resources were re-directed to fighting the pandemic. However, there were dengue cases reported in the country towards the end of 2022 and in 2023 [5]. This outbreaks also coincided with expanding reach of dengue which has caused increased burden globally.

The recurring outbreaks of dengue in Kenya since its re-emergence in 2010 raises questions around whether the cases are linked to the ongoing global dengue outbreaks. The two key questions that need to be addressed are 1) are the dengue cases associated with the outbreaks in Kenya seeded into the country through the ports of entry linking Kenya to other countries or 2) are locally circulating strains that periodically cause outbreaks after spill-overs, and subsequent human-human transmission mediated by mosquito vectors. In this study, we undertook an active investigation of the 2023 dengue outbreak in Mombasa to understand the link between the local dengue outbreak cases to the previous cases and outbreaks as well as the global explosive outbreaks of dengue that have been reported in several countries over the recent past.

## Methodology

### Ethics statement

This study was conducted using anonymized human serum samples obtained through arbovirus surveillance activities and a dengue outbreak investigation study carried out in 2023. Ethical approval for the use of these samples was obtained from the relevant institutional review boards at Kenya Medical Research Institute (KEMRI) and Technical University of Mombasa (TUM). The approval to use surveillance samples at KEMRI was granted by KEMRI’s Scientific Ethics Review Unit (SERU #3035) and approval to use samples that were collected as part of ongoing surveillance studies at TUM received ethical clearance from the Ethical Review Committee of TUM (EXT/005/2022). In addition, samples collected during the active outbreak response in 2023 received ethical clearance from KEMRI-SERU (SERU #4499).

### Sample collection and detection of DENV

The primary set of samples for this study was collected during the 2023 dengue outbreak in Mombasa, Kenya (4.0435° S, 39.6682° E) (Figure 1). A total of 406 blood samples were collected between March and July 2023 from patients presenting with symptoms consistent with arboviral infection. To contextualize the 2023 outbreak within the broader pattern of dengue virus (DENV) activity in the region, we included additional DENV-positive samples previously collected through ongoing surveillance efforts. These included samples from the KEMRI and TUM. The KEMRI samples were collected as part of routine arbovirus surveillance conducted by the Kenyan Ministry of Health through the Division of Disease Surveillance and Response (DDSR). Through this system, local health officials identify suspected arboviral cases based on symptoms consistent with arbovirus infection, such as acute febrile illness, headache, muscle or joint pains. Blood samples are then collected and referred to KEMRI for diagnostic testing. Samples from TUM were part of ongoing surveillance studies tracking febrile illness cases in Mombasa and Kwale Counties along the Kenyan coast. The additional set of samples included 11 samples collected between 2020 and 2022, and 4 more obtained in 2024 and 2025 (Table 1).Serum samples were collected from patients of all ages who presented at local health facilities with symptoms consistent with arboviral infection. The blood samples were immediately stored in liquid nitrogen tanks, pending transportation to the testing laboratories located at KEMRI and TUM. To detect DENV RNA and assess cycle threshold (Ct) values, viral RNA was first extracted using QIAmp Viral RNA Mini Kit (Qiagen, Germany). The extracted RNA was then used to screen for arboviruses using primers targeting the Flavivirus genus [11]. Flavivirus-positive samples were then tested with dengue specific primers, including serotype-specific primers for DEN-1 through DENV-4 [12].

**Table 1:** Demographic and clinical characteristics of the DENV positive cases (n=56) that were analyzed in the study.

| Age range(y)/Sex |  | Counts | % |
| --- | --- | --- | --- |
| 0-5 | F | 2 | 3.64 |
|  | M | 4 | 7.27 |
| 6-12 | F | 1 | 1.82 |
|  | M | 1 | 1.82 |
| 13-19 | F | 4 | 7.27 |
|  | M | 5 | 9.09 |
| 20-44 | F | 20 | 36.36 |
|  | M | 16 | 29.09 |
| 45+ | F | 1 | 1.82 |
|  | M | 1 | 1.82 |
| **Clinical symptoms<sup>a</sup>** |  |  |  |
| Headache |  | 47 | 83.9 |
| Joint/ Muscle pain |  | 46 | 82.1 |
| Fever(>38°C) |  | 21 | 37.5 |
| Abdominal pain |  | 18 | 32.1 |
| Vomiting |  | 15 | 26.8 |
| Epigastric pain |  | 11 | 19.6 |
| Diarrhoea |  | 6 | 10.7 |
| Haemorrhage |  | 4 | 7.1 |
| Low urine output |  | 1 | 1.8 |
| **Serotypes** |  |  |  |
| DENV-Unknown |  | 35 | 57.1 |
| DENV-1 |  | 10 | 17.9 |
| DENV-2 |  | 11 | 19.6 |
| DENV-3 |  | 3 | 5.4 |
| **Number of Samples sequenced <sup>b</sup>** |  |  |  |
| 2020 |  | 2 | 5.3 |
| 2021 |  | 2 | 5.3 |
| 2022 |  | 7 | 18.4 |
| 2023 |  | 24 | 63.2 |
| 2024 |  | 2 | 5.3 |
| 2025 |  | 1 | 2.6 |

<sup>a</sup> This includes information reported by participants during the visit to the clinic.
<sup>b</sup> Includes samples from the 2023 outbreak and contextual samples selected from other years

**Figure 1:**
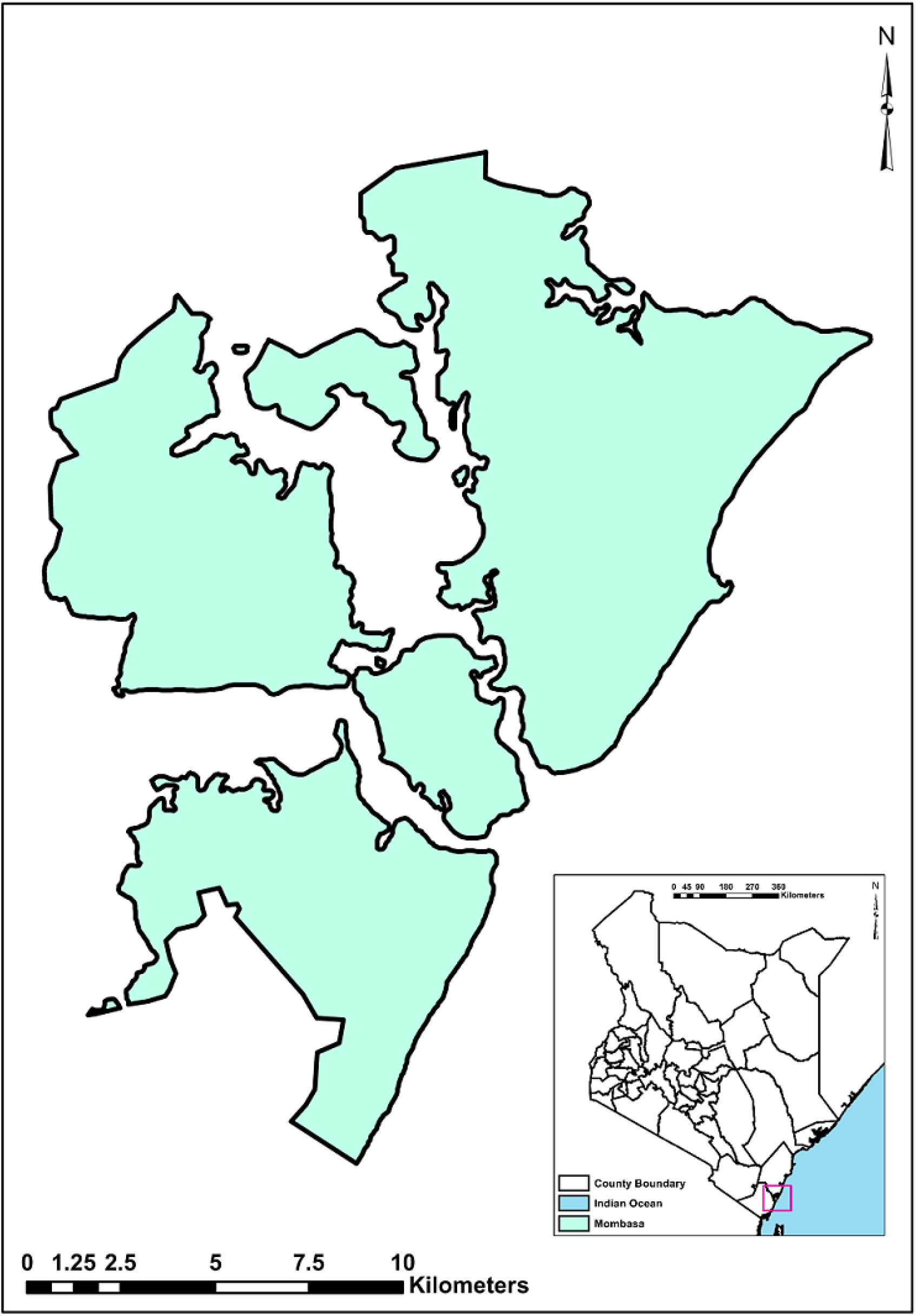
Map of the county of Mombasa where the outbreak investigation was conducted in 2023. The location of Mombasa within the larger map of Kenya is indicated in the in-set (pink rectangular).

### Library Preparation and Sequencing

The extracted RNA was first subjected to DNase digestion using the Turbo DNase kit (Ambion, US) following the manufacturer’s recommendations. Cleaned RNA was used as input for library preparation using NEBNext Ultra II RNA Library Prep Kit for Illumina (NEB, UK). We used the manufacturer’s recommended protocol which was modified to incorporate human rRNA depletion using Qiagen rRNA FastSelect kit (Qiagen, US) and addition of External RNA Controls Consortium (ERCC) controls (Thermofisher, US). The prepared libraries were sequenced on an Illumina iSeq100 and a NextSeq550 system. Raw fastq sequence reads were analyzed in CZ ID pipeline v8.3.15 (https://czid.org/) to remove host and low quality reads as well as assembly and consensus genome generation. Water controls included during library preparation were used to create a mass-normalized background model that was subsequently used to filter background signal. The CZ ID consensus genome pipeline v3.5.0 was used to create contiguous genomes based on the respective dengue serotype reference genomes obtained from the NCBI database (DENV-1: MN923102.1, DENV-2: MN577561.1, DENV-3: JN662391.1).

### Phylogenetic Analysis

The newly generated genome sequences were first genotyped using the DENV typing tool available in Genome Detective (https://www.genomedetective.com/app/typingtool/dengue/). Phylogenetic analysis was conducted using the Augur bioinformatics toolkit [13]. To obtain the dataset for use in phylogenetic analysis, we downloaded all available complete and near-complete DENV genomes (9600–11700 kb) from GenBank (with collection dates up to December 31, 2021) and from GISAID (with collection dates between January 1, 2022, and March 31, 2025). Genomic sequences collected since 2022 were obtained exclusively from GISAID database, due to the predominance of recent entries, particularly those from Africa, being available through this repository. Only genomes with accompanying metadata information including the year of collection and country of origin were included in the analysis. The downloaded set of sequences were combined with the genomes obtained from the current study. Subsequently, the sequences were grouped by country and year of collection and a subset of a maximum of 12 genomes were randomly selected from each group so as to minimize geographical sampling bias. This down sampling resulted in a total of 2332, 2570 and 658 high quality genomes of DENV-1, DENV-2 and DENV-3, respectively. Multiple sequence alignments and phylogenetic inference were performed with the Augur toolkit using MAFFT [14] and IQ-TREE, with the Generalized Time Reversible (GTR) model [15]. The resulting raw phylogenies were further processed using TreeTime [22] to obtain a time-calibrated phylogeny as well as to infer the ancestral traits of the tree nodes, including amino acid changes and geographical location.

## Results

### The circulating DENV cases

In the study, we analyzed a total of 406 serum samples from suspected cases of dengue collected during the 2023 outbreak in Mombasa (Figure 1). Of these, 56 were confirmed to be positive for DENV by qRT-PCR. Three dengue serotypes including DENV-1, DENV-2, and DENV-3 were identified during the outbreak. Among these, the majority were attributed to DENV-1 (10 cases) and DENV-2 (11 cases), while a smaller number were caused by DENV3 (3 cases) (Table 1). Generally, the most affected age group were adults between the ages of 20 and 44 years old. This group accounted for more than 65% of all the cases detected in the study (Table 1). Majority of the cases were associated with headache, joint and muscle pains as well as high fever among other symptoms. Hemorrhagic manifestations were observed in four cases, two of which were confirmed to be DENV-1 (Table 1).

### mNGS Sequencing and Phylogenetic characterization

We sequenced 24 samples associated with DENV cases from the 2023 outbreak in Mombasa. To provide broader context for the outbreak, we also included 14 previously confirmed DENV-positive samples collected between 2020 and 2025. In total, 38 DENV samples with low Ct (<30) values were sequenced using a metagenomics next-generation sequencing (mNGS) approach. The dataset comprised samples from multiple locations within the country including Mombasa, Kwale, Lamu, Garissa and Wajir. Among the sequenced genomes, we identified 21 DENV-1 genomes, 14 DENV-2 and 3 DENV-3. Genotyping analysis classified all sequences into single genotypes for each serotype: Genotype III for DENV-1, genotype II for DENV-2 and genotype III for DENV-Phylogenetic analysis showed that the circulating DENV-1 strains formed a single cluster within lineage A.2 in genotype III (Figure 2A). This strain has been detected in Kenya since 2020, and it shares a common ancestor with strains from other countries in Africa (Figure 2A). Similar strains were previously identified in several Eastern African countries prior to 2020 and in West Africa since 2017. The West African clade has a distinct clustering pattern that differentiates it from the Eastern African clade. However, they both share a common ancestor which is estimated to have existed in the continent around September, 2014 (Figure 2A). Phylogenetic analysis of DENV-2 revealed that the circulating strains belong to lineage A.2 of genotype II. The current strain formed a ladder-like cluster with the earlier strains that were linked to dengue outbreaks that occurred between 2010 and 2017 in different parts of Kenya (Figure 3A). In addition, our analysis showed that the 2023 outbreak strain evolved from these earlier strains. Interestingly, the current circulating strain shares a common ancestor with a strain detected in China and Thailand in 2023 (Figure 3A). Our analysis suggests that this common ancestor existed in Kenya at around October 2023 (Confidence interval: 2020-05-16, 2021-04-27) (Figure 3A). For DENV-3, three sequences were recovered from the 2023 outbreak and our analysis showed that they form a similar cluster within lineage B in genotype III (Figure 4A). Although distinct, the 2023 outbreak strain clustered closely with strains that were responsible for the 2023 dengue outbreak in Ethiopia, a neighboring country to the north. In addition, a related isolate was detected in Italy in August 2023 and it was reported to have been imported from East Africa during the same period [19]. Analysis of the mutational pattern associated with the Kenyan outbreak revealed that DENV-1 had three signature mutations on NS1: V89I, NS4A L19Q and one on NS5: S810T (Figure 2B). On the other hand, DENV-2 had 7 unique mutations: 3 in the NS5 gene and one mutation each in the E, NS1, NS2A and NS4B genes (Figure 3B). DENV-3 strains had two mutations in the E gene and one in the NS4B gene (Figure 4B).

**Figure 2:**
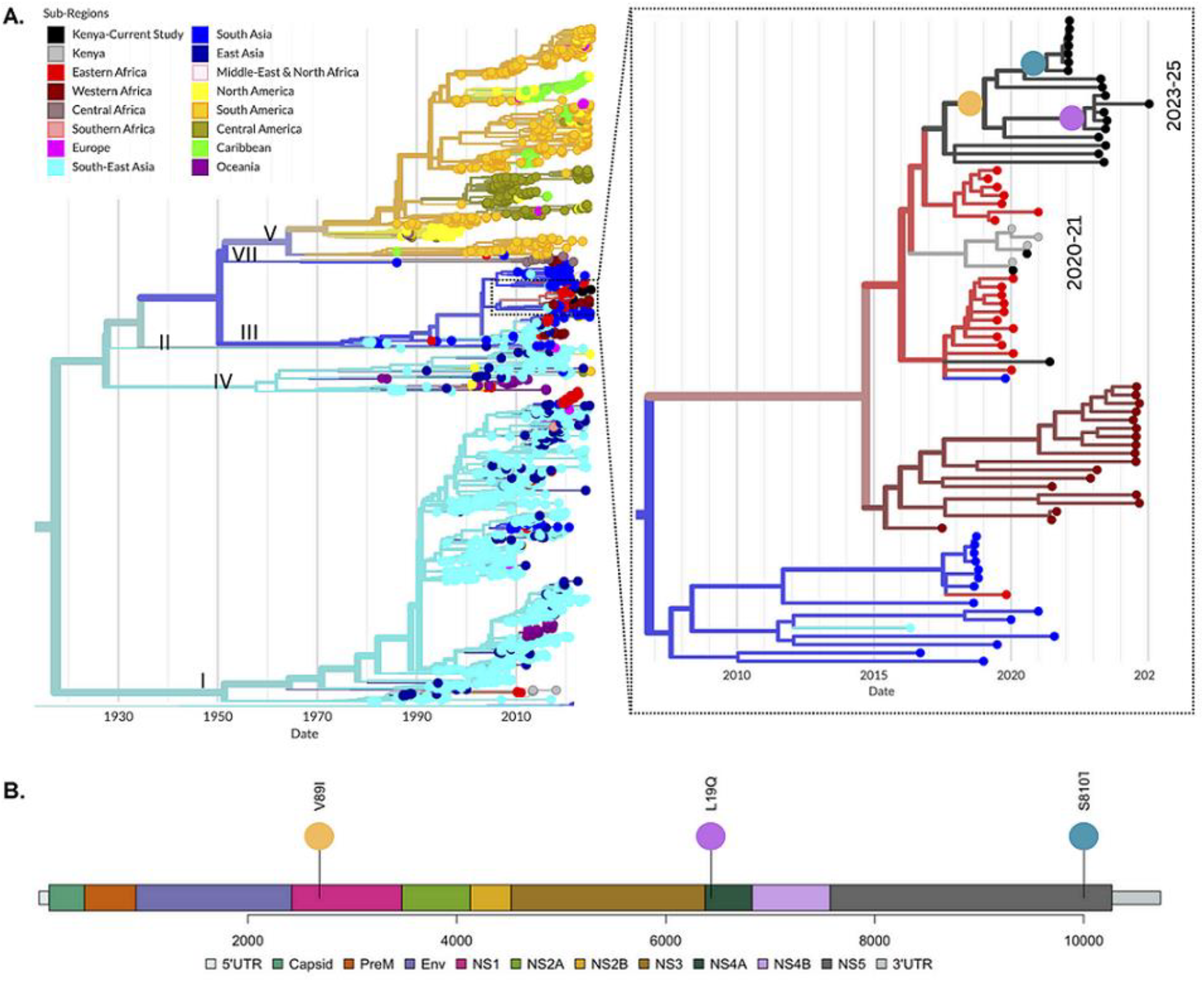
A) Time-resolved maximum likelihood phylogeny of DENV-1 including strains obtained from the study and reference sequences sampled worldwide. The tree branches have been colored according to the geographic origin of the sequences. The inset on the right represents an expansion of the branch containing strains from the study as well as closely related strains from other countries. B) Genome map showing amino acid mutations common among the DENV-1 strains generated in the study. The position in the phylogeny where the mutations occur are indicated in the inset in A, with the color of the dots in the tree branches corresponding to the color of the lollipop depicting the mutation.

**Figure 3:**
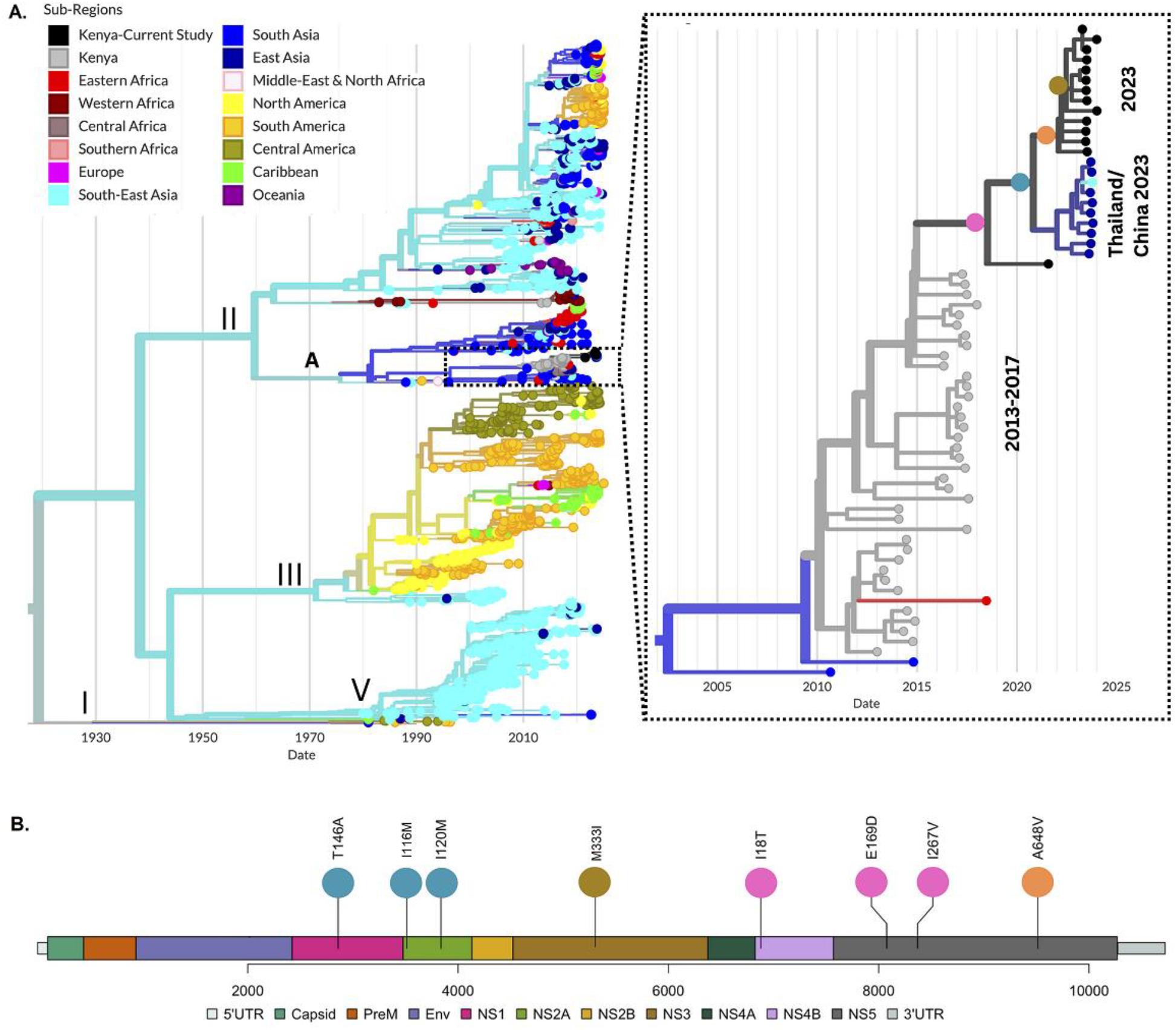
A) Time-resolved maximum likelihood phylogeny of DENV-2 which includes strains obtained from the study and reference sequences sampled worldwide. The tree branches are colored according to the geographic origin of the sequences and the inset on the right represents an expansion of the branch containing strains from the study as well as closely related strains from other countries. B) Genome map showing amino acid mutations common among the DENV-1 strains generated in the study. The position in the phylogeny where the mutations occur are indicated in the inset in A, with the color of the dots in the tree branches corresponding to the color of the lollipop depicting the mutation.

**Figure 4:**
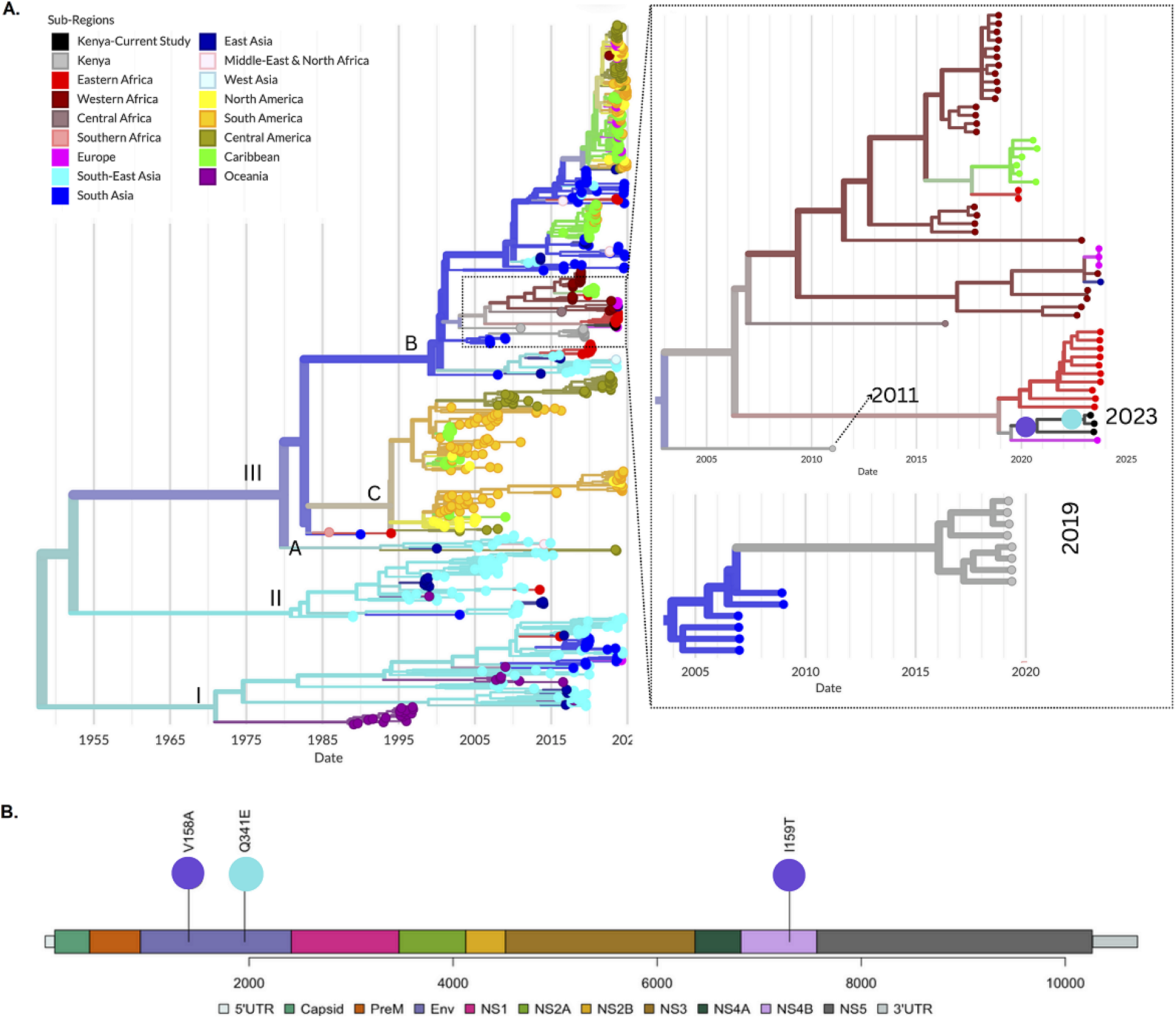
A) Time-resolved maximum likelihood phylogeny of DENV-3 including strains obtained from the study and reference sequences sampled worldwide. The tree branches are colored according to the geographic origin of the sequences and the inset on the right represents an expansion of the branch containing strains from the study as well as closely related strains from other countries. B) Genome map showing amino acid mutations common among the DENV-1 strains generated in the study. The position in the phylogeny where the mutations occur are indicated in the inset in A, with the color of the dots in the tree branches corresponding to the color of the lollipop depicting the mutation.

## Discussion

In this study, we conducted a genomic investigation of the 2023 dengue outbreak that occurred in Mombasa County, with the aim of exploring the relationship between locally circulating DENV strains and the strains responsible for the global dengue emergence. Our investigation confirmed co-circulation of three serotypes of dengue (DENV-1, DENV-2, and DENV-3) during the 2023 outbreak. Our findings also revealed that the majority of cases occurred among adults aged between 20 and 44 years. This demographic group has a higher risk of exposure due to increased daytime activity, which corresponds with the biting behavior of the main mosquito, *Aedes aegypti*. Our findings also show that most of the cases were associated with headaches, joint and muscle pains as well as high fever. Notably, hemorrhagic manifestations were observed in four cases, likely due to secondary heterotypic DENV infection which is a phenomenon commonly observed in regions with multiple co-circulating serotypes [21]. These findings underscore the risk of severe dengue, including Dengue Hemorrhagic Fever (DHF) and Dengue Shock Syndrome (DSS), which are associated with elevated case fatality rates [20, 21].

Phylogenetic analysis of DENV-1 revealed that the circulating strains in 2023 belonged to A.2 lineage. The same strain was previously linked to dengue outbreak in Mandera and Wajir in 2022 as well as Lamu and Mombasa in 2020. This suggests multi-year circulation of this strain across the coastal and north-eastern regions since at least 2020, when the earliest case existed in Kenya. This Kenyan strain shares a common ancestor with strains circulating in other Eastern African countries including Tanzania as well as Mayotte and Reunion Islands [27]. Similarly, this strain also shares a common ancestor with Western African strains circulating in Nigeria, Benin, Burkina-Faso and Togo [28, 29]. This strain appears to be unique to the African continent and has been detected locally since 2017, having diverged from the South Asian strains detected mostly in India at around 2006 (Figure 2A). The findings in this study, therefore, demonstrate sustained local and regional circulation of this strain, with confirmed outbreaks in several African countries including Kenya [27, 28, 29].

The DENV-2 phylogenetic analysis revealed the circulation of one lineage A.2 of genotype II. This particular strain was first introduced into coastal Kenya around 2008 [16] and our analysis suggests it has been circulating within the country ever since. There have been no detection of this strain outside Kenya until around April 2023 when it was detected during an outbreak in Guangzhou, China [23]. The outbreak in China was linked to several imported cases that subsequently led to an epidemic [24, 25]. Phylogenetic analysis reveals the China 2023 and Kenyan 2023 outbreak strains share a common ancestor, which was likely imported into China from Kenya towards the end of 2021 (Figure 3A). Further, the clustering pattern suggests these 2023 strains evolved from earlier Kenyan outbreak strains, 2010-2017, but the long branch leading to the 2023 strains suggests potential circulation in unsampled outbreaks between 2017 and 2023. Analysis of the mutational pattern of the 2023 outbreak strain found the majority of the changes in NS5 gene. The NS5 gene in DENV plays a crucial role in viral replication and inhibition of the host interferon (IFN) signaling [25]. Therefore, given its potential role in virus-host interaction, it would be crucial to determine the effect of these mutations on the long-term local circulation of this lineage.

The phylogenetic analysis of DENV-3 revealed that the circulating 2023 outbreak strain belonged to lineage B.2 of Genotype III, which was also associated with outbreaks in 2011 and in 2019 [30, 31]. However, the strains detected during each of the outbreaks were genetically distinct. The 2019 outbreak strain circulated mostly in South Asian countries, specifically Pakistan, around 2006 to 2009 [35]. However, phylogenetic analysis suggests that this strain may have circulated in unsampled outbreaks prior to causing the outbreak in Kenya in 2019 [30]. On the other hand, the 2011 strain shares a common ancestor with strains responsible for the Kenyan 2023 outbreak. Further, the Kenyan strain is in a cluster that is dominated by strains detected across the African continent; specifically in countries that include Ethiopia, Senegal, Burkina-Faso, Benin and Gabon. The clustering pattern of this African clade is characterized by long branches, suggesting unsampled diversity of this lineage over a long period. Nonetheless, our analysis points to ongoing regional circulation in sub-Saharan Africa, with recently confirmed outbreak cases in Senegal, Ethiopia, Benin and Burkina-Faso [18, 31-34].

## Conclusion

This study provides a comprehensive genomic characterization of the 2023 DENV outbreak in Coastal Kenya. Our analysis confirmed the co-circulation of three DENV serotypes; DENV-1, DENV-2 and DENV-3. Majority of the confirmed cases were are attributed to DENV-1 and DENV-2, but concurrent circulation of all three serotypes raises significant public health concerns, especially the increased risk of secondary infections by the different serotypes and the potential for severe disease manifestations such as dengue hemorrhagic fever (DHF) and dengue shock syndrome (DSS). Phylogenetic analysis revealed the long-term local circulation of DENV-2 in Kenya, with recent exportations of this Kenyan strain into China. On the other hand, DENV-1 and DENV-3 strains detected during this outbreak are genetically related to strains involved in the 2023-24 multi-country dengue outbreaks reported across Eastern and Western Africa, pointing to regional viral movement and possible travel-associated introductions. Our findings, therefore, call for more integrated strategies to contain the dengue outbreak in Kenya, including measures to curtail the long-term local circulation as well as measures to control travel-related importation of new dengue strains from countries within the region.

## Data Availability

The genome sequences generated in the study are available in GenBank under accession numbers; PV717358-PV717395.

## Acknowledgement

The study was made possible through the support of Kenya Medical Research Institutes Internal Research Grants (KEMRI-IRG) under award number KEMRI/IRG/EC0010 as well as Bill and Melinda Gates Foundation/Chan Zuckerberg Initiative (CZI) grant INV-050635 and INV-046964. The findings and conclusions in the study are those of the author(s) and do not necessarily represent the official position of the funders.

